# Decentralisation of the compliance of anti-tobacco law in India: The case of higher educational institutions in New Delhi, India

**DOI:** 10.1101/2023.06.19.23291587

**Authors:** Raja Singh

## Abstract

This paper studies the decentralised compliance responsibility of India’s tobacco control legislation that the government has outsourced to higher educational institutions through an example of New Delhi. 36 higher educational institutions, including universities, are selected and studied through a unique methodology using India’s transparency law, i.e., the Right to Information Act 2005. The study looks into the three most important parameters of decentralised compliance. Two of these require the installation of signboards by educational institutions, and the third involves imposing and collecting fines against persons found smoking within the educational institutions. Regarding the boards, the first board is about the warning prohibiting the sale within 100 yards of educational institutions, and the second one prohibits smoking in educational institutions. The study also asks the educational institutions whether there is a presence of cigarette and tobacco product vendors within 100 yards of the institution, where the sale of such products has been banned by law. The study also found educational activities to create awareness in the institutions for tobacco control and cessation. The results show that the intent to decentralise the compliance of the tobacco control law in New Delhi has not been universally successful and requires much effort.t for its on-ground penetration. Such studies have a policy impact as they serve as an example for the generalisability of such statutes, which only work when there is implementation from the bottom-up and when the deterrent is also strong with incentivisation to the educational institutions to implement tobacco control with vigour.

## Introduction

India has an anti-smoking and anti-tobacco law that was passed in the year 2003. It is known as the Cigarettes and Other Tobacco Products (Prohibition of Advertisement and Regulation of Trade and Commerce, Production, Supply and Distribution) Act, 2003 or the COTPA Act, 2003 ^1^. The intent of the law was to enact *‘a comprehensive law on tobacco in the public interest and to protect the public health’* and to *‘prohibit the consumption of cigarettes and other tobacco products which are injurious to health with the view of achieving the public health in general as enjoined by Article 47’* of the Indian Constitution ^2^. This was made in response to the 43^rd^ World Health Assembly, where member states were urged to consider tobacco control strategies, with a focus on protecting children from voluntary exposure to tobacco use and discouraging the use to tobacco ^3^. It is this focus on young people that the Indian law focused on educational institutions and minors. The legal manifestation of this concern for minors and educational institution vicinity is seen in section 6 of the COTPA Act, 2003 states that:

> *No person shall sell, offer for sale, or permit sale of, cigarette or any other tobacco product-*
>
> *(a) to any person who is under eighteen years of age, and*
>
> *(b) in an area within a radius of one hundred yards of any educational institution*.

This clearly shows that the intent of protecting young children from tobacco and smoking existed, even if it was in response to a national commitment in front of an international forum. But the most important and missing link in this is the way this would be implemented. The concern was raised even in the parliament when this bill was taken in the Lok Sabha on the date of its adoption as a law passed by the legislature in India after one house of the parliament had already passed it. The concern raised was whether simple enactment of the law will actually lead to its implementation?^4^

This leads us to the second decision that the government made to solve the problem of the implementation of the COTPA Act for its implementation, especially with respect to banning the sale of tobacco products within 100 yards of educational institutions. The government simply outsourced and decentralised the compliance to educational institutions by enacting a rule under section 31 of the COTPA Act called the Cigarettes and Other Tobacco Products (Display of Board by Educational Institutions) Rules, 2009^5^, which states as follows:

> *3. Display of Board by Educational Institutions.(1) The owner or manager or any person in-charge of affairs of the educational institution shall display and exhibit a board at a conspicuous place outside the premises, prominently stating that sale of cigarettes and other tobacco products in an area within a radius of one hundred yards of educational institutions is strictly prohibited and that it is an offense under Section 24 of the Act with fine which may extend to two hundred rupees.’*

By this act the government outsourced and decentralised the compliance to educational institutions and that too with the only requirement of putting up a signboard which prohibits anyone from selling cigarettes and tobacco products within 100 yards of educational institutions. There may also be scope for an assumption that putting such a board may be deterrent enough, as even if someone has been caught, the meagre punishment of a fine of rupees two hundred will not only be too little but will also be over as s summary trial. The lawmakers, while discussing it, also had similar concerns as this may not be a deterrent enough, and people may pay the money as there is no severe punishment for the re-occurrence of the offence by the same person^4^.

Another way by which compliance has been decentralised is by giving two more responsibilities to educational institutions. One is the installation of a board which states *‘No-Smoking Area-Smoking here is an Offense’* under the Prohibition of Smoking in Public Places Rules, 2008^6^, at multiple places, including the entrance, other conspicuous places and one on each floor near the staircase. The second is even the on-ground implementation of the prohibition of smoking in public places by making the College/School/Headmaster/Principal or even the teacher as the authorised officer to impose and collect the fine against the violation of Section 4 of the COTPA Act, 2003. This means the teachers have been expected to be the law enforcers and collect fines from students who are found smoking in educational institutions. These have been reiterated in intent by the guideline of the Delhi State Tobacco Control Cell^7^ to educational institutions and are also in line with the earlier Delhi Assembly statute^8^, which preceded the national anti-tobacco/anti-smoking Act ^1^.

With this, we find out that there are three responsibilities of the educational institutions by which the anti-tobacco law has to be implemented and enforced by them. These are summed up as follows:

1. Putting a poster prohibiting the sale of cigarettes and tobacco products within 100-yard vicinity of educational institutions.
2. Putting a board prohibiting smoking within educational institutions.
3. Imposing and collecting the fine from anyone found smoking in educational institutions.

To check compliance for the above three points, this study has been designed at the level of the higher educational institutions in New Delhi. The aim of this study is to find the decentralised implementation of the COTPA Act 2003 and its rules by educational institutions in New Delhi. This compliance has been checked for national-level institutes by the same author ^9^, but the compliance on the level of Delhi, with respect to fine collection and placement of indoor posters, has not been covered fully by any other study from India or from the world for that matter. The author, in an earlier study, has looked for the checking of the signboard compliance prohibiting the sale of tobacco products within 100 yards of the vicinity but has found non-universal compliance with the signboard requirement ^10^.

The need for this study is justified as this may be the first study looking comprehensively at the compliance by the educational institutions in the heart of India, in its capital city of New Delhi.

## Materials & Methods

This study focuses on higher education institutions. 47 higher education institutions, most of them run by the Government of National Capital Territory of Delhi, have been included. This number is of randomly selected higher educational institutions, including 4 universities run by the Government of NCT of Delhi located in New Delhi and all 12 colleges of the Delhi University that are fully sponsored by the Delhi government, i.e., colleges funded 100% by the Delhi Government, but under the Delhi University^11^. The main campus of the Delhi University has also been included. Many colleges of the universities and other autonomous stand-alone colleges run directly by the Delhi Government have been included. This is a fairly representative sample, and the knowledge of the exact number of all the colleges run by various governments and the lack of calculation of sample size may be a limitation of the study. But covering 4 universities, all Delhi government-run colleges of the Delhi University provide a fairly good representation of higher educational institutions in New Delhi. The author has performed a separate study for the colleges run by the Central Government in India, which also includes the centrally run colleges in New Delhi like the Indian Institute of Technology, New Delhi.

This study uses a unique methodology, where the transparency law of India has been used for data collection from the various institutions that have been included in the study. The Right to Information Act of 2005, usually otherwise used for holding the government to account, has also found its use in this study where it has been used for research^12,13^. This is just like the FOIA or the Freedom of Information Act of the United States. Out of the 47, 11 institutions did not respond to the request for the information in defiance of the Right to Information Act 2005. This can lead to penalty and disciplinary action, and the author has invoked Section 19 and will be following up with the institutions as they have not responded to the request, which is not only prohibitive for research but is also illegal as per law. In this study, therefore, 36 institutions have finally been included out of the 7 that were reached out.

The information asked for was in the sense of checking the decentralised application of the COTPA Act 2003 by the educational institutes.

The information was asked in the form of applications filed under Section 6 of the Right to Information Act 2005. This mandates the public authority, which in our case is the educational institutions run by the Government of NCT of Delhi as mentioned before, to provide the information under the seal and signature of a senior officer of the public authority, which is usually a senior faculty in the case of educational institutions. This information has to be supplied within a mandatory time period of 30 days as per Section 7(1) of the Right to Information Act 2005. This approach enables only authentic, duly certified information and makes the information accurate. This information, since released under the public transparency law, also becomes information in the public domain and is available for wide public usage. A limitation of using the Right to Information Act 2005 is twofold. Firstly, the information has not been ground-truthed by the author, and it relies on the information provided by the officer. But since the information is of legal nature, this apprehension may be diluted as the government officer will not provide false information under their sign and seal. Secondly, another limitation is that this approach is only valid for educational institutions which come under the category of public authority as per Section 2(h) of the RTI Act 2005. This automatically excluded the private educational institutions that are present in New Delhi but are not included in this study. Due to the exclusion of these under the transparency law, these private institutions become somewhat of opaque and cannot be mandated by law to provide information of public interest, and researchers may need to depend on the choice and will of these institutions to provide such information, and this may lead to the provision of no information at all. This is because, due to the absence of a legal requirement, information which may put the institution in a bad light may never be available for researchers, journalists and others interested.

The educational institutions, once selected, were sent an application as mentioned before with the following point of information asked:

1. Information regarding the presence of the signboard on the outside of the educational institution in compliance with the Cigarettes and Other Tobacco Products (Display of Boards by Educational Institutions) Rules, 2009^5^, which instruct the educational institutions to put up a board which states to prohibit the sale of cigarettes and tobacco products in an area within a radius of one hundred yards.
2. Information regarding the presence of boards and their location state that the educational institution is ‘No Smoking Area-Smoking Here is an offence’ as per Section 3(b) and Schedule II of the Prohibition of Smoking in Public Places Rules, 2008^6^.
3. Information of the total instances of fines collected/offences compounded/offences recorded/warning issued in respect to Section 5 of the Prohibition of Smoking in Public Places Rules, 2008, which stated the authorised officers who are competent to act under Schedule III of the Prohibition of Smoking Rules, 2008 from the year 2012 till the year 2022. In this, the Principal/teacher/director/head of the institution is a person authorised to take action under the law.
4. Information on the presence and number of vendors of cigarettes and other tobacco products within a distance of 100 yards from the outer boundary of the educational institute.
5. Information regarding the list of events/initiatives/activities/circulars/anything on record where the educational institution has taken steps to prevent use of cigarettes and tobacco products.

An annexure with the detailed quoted laws was also annexed with the application.

The institutions provided the reply through the RTI portal^14^ as well as written replies with signatures on the letterhead of the institution. The period of collection of the information has been between April to May 2023. The information from the educational institutions was received in three ways. Some sent a detailed hard copy; others uploaded it to the Right to Information portal and some sent it via email. This information was collected till the end of the one-month information period and even extended by another month of grace period to cater for the postal delays or any other delays.

These results were compiled in the form of a table, and then the results were reported, and interpretation was drawn and made in line with the aims and objectives of the study.

Another limitation that is reported is that the first reply by the educational institutions has been reported, and the first appeal or the second appeal has not been filed to obtain clarification on the information provided in the first reply by the educational institutions.

### Non-requirement of ethics review for this study

This study uses data available in the public domain that has been supplied by a public authority under the Right to Information Act 2005. There is no involvement of any human participant, human subject, human or animal tissue. There is also no linked identifier to any human participant or subject. The mere fact that the data has been supplied under the open public domain through the Right to Information Act 2005 deems it non-personal as, according to the transparency law, the public authority cannot provide any third-party information by law.

## Results

In general, out of 36 institutions that provided the information, on the issue of the provision of the signboard prohibiting the sale of cigarettes and tobacco products within a vicinity of 100 yards, 16 have provided the board in the format required, which means the board stated explicitly that the ‘Sale of cigarettes and other tobacco products within 100 yards of the educational institutions is an offense…’. 7 stated clearly that the board has not been provided, though, from the reply, many of the 7 were in the belief that the placement of the board was not their own responsibility but was of some other agency. This will be discussed in the paper later. 7 out of the 36 stated there was a presence of the board, but it was not clear whether the format was as required, as many of these 7 stated that they had placed ‘No Smoking’ boards instead of the boards which prohibit the sale within 100 yards vicinity. It is also pertinent to note that 6 out of the 36 that provided information in general did not provide the information related to the signboard prohibiting sale within 100 yards. They either stated that the information is either not on record or used some other way of denying the information.

But, with respect to the placement, the simple signboards prohibiting smoking in educational institutes as public places, the signboard compliance was more promising. Out of 36 institutions, 32 have the boards placed. 2 institutions did not place the boards, and 2 did not provide the information.

On the matter of the recording instances of smoking of instances of collecting the fines, these two were together clubbed, and it was found that only 1 institution has a record of smoking instances/fine collection. 17 educational institutions had no instances of smoking recorded/no fines collected. For this, a staggering 18 institutions did not provide the information or did not have such information on record.

With respect to the issue of whether there was a presence of tobacco vendors within 100 yards of the educational institution vicinity, it was found that 2 institutions reported the presence of vendors. 17 on the other hand, stated that there was no presence of the vendors. 7 institutions responded that this information was either not under the purview of the educational institution or was not a responsibility of the educational institution. Some even went ahead and transferred this part of the information either to the Delhi Police or to the health department. 10 educational institutions denied the information or did not provide or stated that the information was not on record.

As education is the major role played by educational institutions in society, we checked the role played by educational institutions in educating about tobacco cessation. We found that out of 36 educational institutions, 27 stated that they conducted activities related to educating students on tobacco cessation. These activities may range from counselling to plays, dramas, talks, and other seminars. One institution reported no activities on tobacco cessation, and 8 did not provide any information and stated that no such information was on record.

## Discussion

This study is not the first one to check the compliance of the COTPA Act 2003 in India, yet it is unique and brings forth new knowledge in new ways. The basis of the study has been the decentralisation of the compliance of the COTPA Act 2003 in educational institutions of New Delhi, India which forms the heart of the second most populated country in the world. The first contribution of this study is that it is the first one which checks the decentralised compliance of educational institutions in Delhi with respect to the detail of the smoking instances checked or fines collected. This is something that is part of the law, where the decentralised duty is given to the principal, headmaster and faculty of the educational institution, but this check is not part of the standard check that forms part of the ‘Self-Evaluation Scorecard for Tobacco Free Educational Institution’ released by the Ministry of Health and Family Welfare, Government of India ^15^. There are also other studies that are related to compliance with the COTPA Act 2003 in India. The first study is which only simply measured advertisements at vendor’s points and only selected 30 schools randomly ^16^. This study did not include signboard compliance or fines compliance. Another group performed two studies performed cross-sectional and proximity analysis^17^, and earlier did a multi-stage cluster sampling survey for checking the advertisement by tobacco vendors^18^. In these, the signboard component has been missed. Another study used questionnaires for compliance. But, even this study was restricted to the ‘No Smoking boards’ compliance and did not include the signboard prohibiting sale in 100 yards vicinity^19^. Another study performed concentrated on tobacco vendors on various parameters, including advertisement and selling the products to minors ^20^. This current study is useful as it deals with three important points related to two formats of signboard compliance and the third being the compliance of smoking instance/fine collection recording. The author has himself performed a similar study on national-level educational institutions and also performed ground truthing of the signboard compliance (the board banning sale within 100 yards) and the actual presence of the vendors within 100 yards of the educational institutions ^10^. Another unique component of this study is the sourcing of the data right from the source itself. Using the Transparency law of India, i.e., the Right to Information Act 2005, ensures that the information provided is most authentic as it has been provided under the sign and seal of the educational institution under a legal requirement, with penal punishments available for the provision of wrong information.

There is also the important issue of the seeming confusion regarding the format of the boards. There were 7 institutions that did not provide the appropriate reply to the issue of the board prohibiting the sale of cigarettes and tobacco products within 100 yards of the educational institutions. Some institutions seemed to be using the two boards interchangeably, where the ‘No smoking’ boards were only represented as the board which prohibited the sale of cigarettes within 100-yard proximity. This has also been confirmed by a ground-truthing study by the author in Delhi.

Another issue of important concern is the belief that some educational institutions had regarding the two issues, where the educational institutions assumed that the duty did not lie with the educational institution but with some external agency like the police or the health department of the government. The first is the placement of the board prohibiting the sale of tobacco within 100 yards of the educational institutions, where the educational institutions in question, stated that this was the responsibility of the police or the government agencies to place the boards. This belief is not only false but is also illegal and in direct contravention to the COTPA Act 2003 and its related rules. Another important factor that the educational institutions believed are not ‘under the purview’ of the educational institution is the presence of cigarettes and tobacco products vendors within 100 yards of the educational institution’s vicinity. This may appear true in the first instance, but from the point of view of decentralisation, it may then be an educational institute that may either be the direct fine imposing authority or may be the first responder by informing the law enforcement agencies. But, from the point of view of the guidelines issued by the Ministry, as stated before, one of the important criteria for self-evaluation for check compliance of the COTPA Act 2003 is the check for the presence of tobacco vendors within proximity of 100 yards. By the presence of this factor in a ministry-issued guideline, the educational institutions cannot absolve themselves of the responsibility of checking for the vendors selling cigarettes and tobacco products and reporting the same to the law enforcement agencies. This has not been followed by the educational institutions that have in the information provided seemingly outsourced this responsibility to either the police or the health department.

As another peripheral issue, the awareness activities organised by the institutions are also an important step by the educational institutions with 75% of the institutions undertaking such activities.

Another important issue mentioned earlier, deals with the collection of fines. The empowerment of the headmaster, principal/head of the institute, or even the faculty member to impose and collect the fines is a decentralised approach to the compliance of the COTPA Act 2003 by the government, made in response to the legislature’s wish of implementing the act. At the first glance, it may appear as a very positive step as the power has been transferred to the actual person on the ground till the last mile, but at the same time, it appears that there may also be concerns regarding the outsourcing of responsibility to the educational institutions.

From the generalisability point of view, it is important to understand that making a statute and effecting its implementation are important points to consider. There may be other such statutes which prohibit speed limits, fast food sale, or any other prohibition which may be top-down in nature. For the statute to actually be implemented, capacity buildings, training, and regular outreach must be part of the process in order to make the implementation more bottoms up in nature.

## Conclusions

The aim of this study is to find the decentralised implementation of the COTPA Act 2003 and its rules by educational institutions in New Delhi. As far as the installation of boards prohibiting the sale of cigarettes and other tobacco products is concerned, the implementation is not universal, with only 44.4% of the institutions having a board. In the case of the no-smoking board, the compliance was better, with 88.9 per cent of the institutions having such boards. On instances of smoking and fine collection, only one institution reported an instance and collected a fine, while per cent of institutions did not have any instance of smoking/collected fine. It is negative that about 505 institutions did not even have a record of this collection of fines and recording of the smoking instances, which can safely be assumed as defiance of the law. In New Delhi, with the sample size studied, it can be stated that compliance with the COTPA Act 2003 is not universal in the complete sense, and there are many areas that need attention. The COTPA Act 2003 must be made stricter with more severe and deterring punishments for the offenders. The police should coordinate with educational institutions by taking the lead and initiative so that the vendors selling cigarettes and tobacco products within 100 yards can be punished severely.

The intent of decentralising the role to the educational institutions has not been universally implemented, with some educational institutions not even owning up to the fact that the compliance of installing a signboard prohibiting the sale of cigarettes in the vicinity of 100 yards falls in their responsibility. A push needs to be made to incentivise educational institutions in the form of academic accreditations and enhanced academic rankings if they are able to fully implement the COTPA Act 2003 in and around their premises.

There is also scope for further study in multiple areas, including the implementation check in all geographies. Another issue is research on whether 100 yards limit is effective and whether this needs to be increased, and whether the simple position of a signboard by an educational institution is deterrent enough for the vendors to not sell cigarettes and tobacco within the vicinity of educational institutions. Also, to check whether the process of imposing fines is an effective measure as a person affording a cigarette may pay the fine and carry on with smoking due to lack of stricter punishments or on recurrence of the offence.

## Data Availability

All data produced in the present study are available upon reasonable request to the authors

## Acknowledgements

Special thanks to Rajshekhar Pullabhatla and Group Captain P Aanand Naidu (retd.) of the Information Sharing and Analysis Center and Tathatara Foundation, respectively. Many thanks to the staff and faculty of the Department of Architecture. Thanks to Shreya G and Praveen K for their support. Many thanks to the Public Information officers of all the institutes for their kind support.

## Declarations

The author declares no competing interests.

This study does not require ethics approval as stated in the ‘Materials and Methods’ above in detail.

## Notes

### Competing Interest Statement

The authors have declared no competing interest.

### Funding Statement

This study did not receive any funding

## References

1. Government of India. The Cigarettes and Other Tobacco Products (Prohibition of Advertisement and Regulation of Trade and Commerce, Production, Supply and Distribution) Act, 2003.; 2003. Accessed February 14, 2023. https://legislative.gov.in/sites/default/files/A2003-34.pdf

2. Bakshi PM. The Constitution of India; Selective Comments. Universal Law Publishing

3. Forty-Third World Health Assembly: Resolutions and Decisions Annexes. World Health Organisation; 1990:15. Accessed June 19, 2023. https://apps.who.int/iris/bitstream/handle/10665/173422/WHA43_1990-REC-1_eng.pdf?sequence=1&isAllowed=y

4. Lok Sabha Debates (English Version) Twelfth Session (Thirteenth Lok Sabha). Lok Sabha Secretariat, New Delhi; 2003:358. Accessed June 19, 2023. https://eparlib.nic.in/bitstream/123456789/759727/1/lsd_13_12_30-04-2003.pdf

5. Ministry of Health and Family Welfare, Government of India. Cigarettes and Other Tobacco Products (Display of Boards by Educational Institutions) Rules, 2009. Accessed February 14, 2023. https://nhm.gov.in/cota/Notifications%20on%20Section-6%28b%29

6. Government of India. The Prohibition of Smoking in Public Place Rules, 2008.; 2008. Accessed February 14, 2023. https://nhm.gov.in/cota/Notifications%20on%20Section%204%20of%20the%20Act%20Related%20to%20Prohibition%20of%20Smoking%20in%20Public%20Places/GSR-417-D%28E%29.pdf

7. Guideline for Tobacco Free schools/Educational Intitutions. Published online December 8, 2009. Accessed June 19, 2023. https://www.edudel.nic.in//upload_2013_14/11008_dt_24072013/3971_76_dt_07012010.pdf

8. The Delhi Prohibition of Smoking and Non-Smokers Health Protection Act 1996. Accessed January 2, 2023. http://www.bareactslive.com/Del/dl155.htm

9. Singh R. Compliance with the Provisions Related to Higher Educational Institutes of Anti-Tobacco/Smoking Law by Institutes of National Importance in India. Health Policy; 2023. doi:10.1101/2023.02.13.23285898

10. Singh R. Signboards prohibiting tobacco sale within 100 yards of educational institutes: the appraisal of prohibition compliance and on-ground status of the anti-smoking law in New Delhi’s major administrative precinct. Cities Health. Published online May 31, 2023:1–10. doi:10.1080/23748834.2023.2215417

11. DU Colleges Sponsored by Govt. of NCT of Delhi | Directorate of Higher Education. Accessed June 19, 2023. https://higheredn.delhi.gov.in/higher-education/du-colleges-sponsored-govt-nct-delhi

12. Singh R. RTI for Research: Using the Right to Information Act, 2005 for Research in India. Vol 1. Sandeep Kaur(BooksBonanza); 2020. 10.5281/zenodo.6088938

13. Republic of India. The Right to Information Act, 2005. Vol 200522.; 2005. Accessed November 17, 2021. https://rti.gov.in/rti-act.pdf

14. RTI Online□:: Online RTI Information System. Accessed June 19, 2023. https://rtionline.delhi.gov.in/

15. Guidelines for Tobacco Free Educational Institution. Ministry of Health and Family Welfare, Government of India Accessed June 19, 2023. https://ntcp.mohfw.gov.in/assets/document/TEFI-Guidelines.pdf

16. Elf JL, Modi B, Stillman F, Dave P, Apelberg B. Tobacco sales and marketing within 100 yards of schools in Ahmedabad City, India. Public Health. 2013;127(5):442–448. doi:10.1016/j.puhe.2013.02.003

17. Mistry R, Pednekar MS, McCarthy WJ, et al. Compliance with point-of-sale tobacco control policies and student tobacco use in Mumbai, India. Tob Control. 2019;28(2):220–226. doi:10.1136/tobaccocontrol-2018-054290

18. Mistry R, Pednekar M, Pimple S, et al. Banning tobacco sales and advertisements near educational institutions may reduce students’ tobacco use risk: evidence from Mumbai, India. Tob Control. 2015;24(e1)e100–e107. doi:10.1136/tobaccocontrol-2012-050819

19. Khargekar NC, Debnath A, Khargekar NR, Shetty P, Khargekar V. Compliance of Cigarettes and Other Tobacco Products Act among Tobacco Vendors, Educational Institutions, and Public Places in Bengaluru City. Indian J Med Paediatr Oncol. 2018;39(04):463–466. doi:10.4103/ijmpo.ijmpo_136_17

20. Goel S, Kumar R, Lal P, et al. How Compliant are Tobacco Vendors to India’s Tobacco Control Legislation on Ban of Advertisments at Point of Sale? A Three Jurisdictions Review. Asian Pac J Cancer Prev. 2015;15(24):10637–10642. doi:10.7314/APJCP.2014.15.24.10637

